# A Deep Learning-based Genome-wide Polygenic Risk Score for Common Diseases Identifies Individuals with Risk

**DOI:** 10.1101/2021.11.17.21265352

**Authors:** Jiajie Peng, Jingyi Li, Ruijiang Han, Yuxian Wang, Lu Han, Jinghao Peng, Tao Wang, Jianye Hao, Xuequn Shang, Zhongyu Wei

**Affiliations:** School of Computer Science, Northwestern Polytechnical University, Xi’an, 710072, China; Key Laboratory of Big Data Storage and Management, Northwestern Polytechnical University, Ministry of Industry and Information Technology, Xi’an, 710072, China; College of Intelligence and Computing, Tianjin University, Tianjin, 300072, China; School of Data Science, Fudan University, Shanghai, 200433, China

**Keywords:** disease risk prediction, deep learning, polygenic risk score

## Abstract

Identifying individuals at high risk in the population is a key public health need. For many common diseases, individual susceptibility may be influenced by genetic variation. Recently, the clinical potential of polygenic risk score (PRS) has attracted widespread attention. However, the performance of traditional methods is limited in fitting capabilities of the linear model and unable to capture the interaction information between single nucleotide polymorphisms (SNPs). To fill this gap, a novel deep-learning-based model named DeepPRS is developed for scoring the risk of common diseases with genome-wide genotype data. Using the UK Biobank dataset, the evaluation shows that DeepPRS performs better than the other two existing state-of-art methods on Alzheimer’s disease, inflammatory bowel disease, type 2 diabetes and breast cancer. Since DeepPRS does not only rely on the addictive effect of risk SNPs, DeepPRS has the chance to identify high-risk individuals even with few known risk SNPs.

## 1. Introduction

One of the main public health needs is to stratify the individuals for a given disease to identify individuals at high risk and enable enhanced screening or preventive therapies^[1]^. For most human diseases, individual susceptibility is influenced by genetic variation to some extent^[2]^. Consequently, one important approach for identifying individuals at high risk is to stratify individuals based on inherited DNA variation^[1]^. According to the number of genes that cause the disease, the diseases involving genetic factors are traditionally divided into single-gene mendelian diseases and complex or common diseases^[3]^. In the 1980s and 1990s, based on linkage analysis and fine mapping within large multiplex pedigrees, efforts to map disease genes mainly focused on rare diseases, monogenic diseases and syndrome-type diseases^[1]^. By 2000, around 1000 single-gene inherited diseases had been characterized, including many diseases that have a significant impact on biomedicine, such as Huntington’s disease^[4][5]^and cystic fibrosis^[6][7]^. However, linkage analysis is very limited for common, later-onset traits with more complex diseases, such as asthma, diabetes and depression^[8]^. Until 2005, genome-wide association studies (GWAS) have identified a large number of genetic variants, mostly single nucleotide polymorphisms (SNPs)^[9]^. The emergence of GWAS provided important clues for the discovery of genetic characteristics that influence the occurrence of complex diseases^[3]^.

In the decades since the first GWAS^[10]^, people’s understanding of the genetic basis of common human diseases has been changed. For complex or common diseases, genetic susceptibility is jointly determined by thousands of mostly common variants, and a single variant has little effect on population risk^[11][12]^. On the basis of GWAS research, polygenic risk score (PRS), which quantifies individual genetic risk, has become a powerful tool for common disease risk prediction^[13]^. PRS quantifies the degree of individual susceptibility to disease by calculating the cumulative effect of multiple susceptibility sites^[14]^. The development of robust PRS for several common diseases has been catalyzed by the continuous expansion of GWAS dataset scale and the establishment of large-scale biobank supporting score verification^[1][2][15][16]^. Many studies have demonstrated the utility of PRS for disease risk stratification as well as their implications for early disease detection, prevention, therapeutic intervention and life planning^[17]^. The traditional PRS method quantifies the impact of variations on individuals based on a simple linear additive model. However, a popular thought suggests that the mechanisms of gene action and the structure of complex traits are actually much more complex than the additive model describes^[18]^. For example, epistasis, defined in functional terms as an event where the influence of one locus depends on the genotype of another, is a type of non-additive genetic variation^[19]^. A study shows that there is an interaction between HLA-C and ERAP1 for psoriasis, and ERAP1 variants only influence psoriasis susceptibility in individuals carrying the HLA-C risk allele^[20]^. Therefore, the traditional additive method of PRS calculation not only fails to utilize the location information of variations, but also ignores the interaction information among various variations. In addition, the traditional PRS model is constructed using classifiers such as logistic regression model^[21]^ and penalty regression model^[22]^. These models have limited model fitting capabilities, which also affects the accuracy of the final prediction results to a certain extent.

Recently, deep learning, a subfield of machine learning, has been successfully applied to several areas, such as medical imaging, health record natural language processing and generalized deep-learning methods for genomics^[23][24]^ ^[25][26]^.

Inspired by the success of deep learning in healthcare area, we hypothesize that deep learning can further enhance the predictive ability of PRS models by integrating large scale genotype data. In this study, we propose a deep learning-based approach to improve the traditional PRS model, and adopt bidirectional long short-term memory network (BiLSTM) as classification model. An efficient disease risk prediction method is presented, named DeepPRS, which can be used to calculate an individual’s polygenic risk score and stratify the population. Here are our three contributions:

- We propose a deep neural network-based model, named DeepPRS, to generate polygenic risk score. Compared with the linear addition of the traditional method, we not only consider the position relationship of SNPs, but also capture the interaction information between long-distance genes on each chromosome through BiLSTM.
- Our risk analysis demonstrated that the disease risk of people with high deep polygenic risk score is much higher than people with low deep polygenic risk score. Therefore, DeepPRS can be used as an early warning sign to initiate disease prevention and screen in high-risk populations.
- Compared with traditional methods, DeepPRS needs less prior information. The performance of DeepPRS is better than the traditional methods, which needs the effect estimation given by GWAS as the genotype weight.

## 2. Result

### 2.1. Overview of the DeepPRS model

DeepPRS combines the genotype data^[27]^, summary statistics results of GWAS^[28][29][30][31]^, and 1000 Genomes reference panel^[32]^ to predict the risk of common diseases (**Figure. 1a, Supplementary Figure. S1** and **Table 1**). In short, we first carry out quality control (QC) on genotype dataset, and then a set of parameters based on GWAS results (*P* values) and linkage disequilibrium reference panel from 1000 Genomes of 503 Europeans (*r*^*2*^) are applied to select SNPs for the downstream analysis. The feature selection of this step provides high-quality and low-dimensional input forgenerating deep polygenic risk score, which can greatly reduce the number of parameters and running time of the neural networks. Then, a novel encoding schema is proposed to represent genotype information. The two dimensions of the feature vector represents risk alleles (Alt allele) and non-risk alleles (Ref allele) respectively. Compared with the traditional additive encoding method, our encoding method can not only express the number of alleles carried, but also avoid additional quantitative relationships, and better deal with missing genotypes (**Figure. 1b**). Finally, the features are fed to deep neural networks for disease risk prediction. We first use a partial connected layer to connect SNPs to genes within 250 kilobases region according to the location information of SNPs, which greatly reduces the feature dimension and effectively reduces the over-fitting phenomenon. Next, we use the BiLSTM layer to capture the interaction information between long-distance genes from the forward and backward directions at the same time, and finally we use the fully connected layer as the classifier. The prediction probability is used as the deep learning-based polygenic risk score (**Figure. 1c**).

**Figure. 1.**
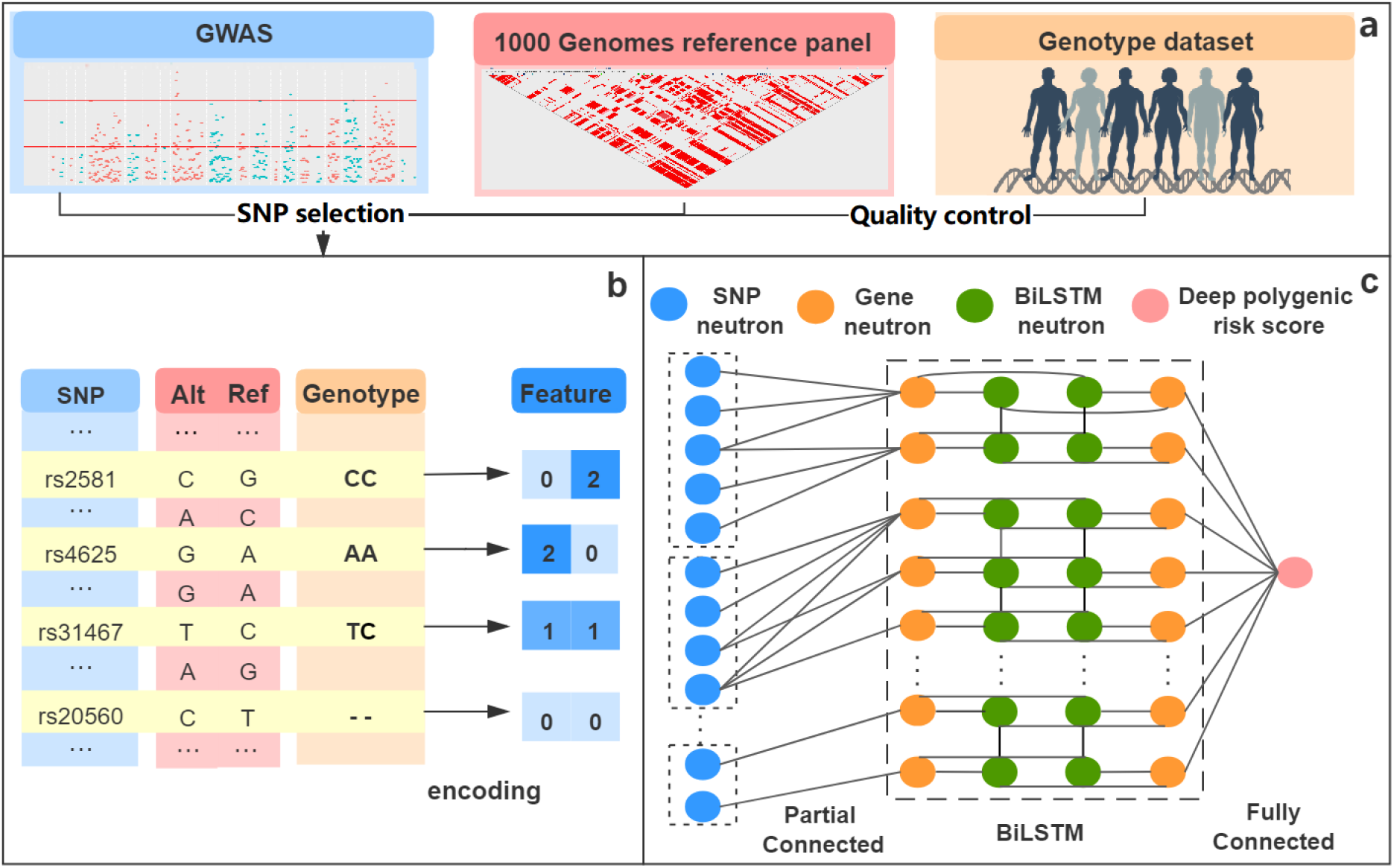
Overview of the DeepPRS method. DeepPRS includes three main components. **(a)**, SNP selection step considers the significance SNPs based on the GWAS result, the linkage disequilibrium reference panel from 1000 Genomes Europeans and the availability of genotype data. **(b)**, The genotype information is encoded into two dimensions of feature vectors, where the first dimension represents the number of non-risk allele (Ref allele), the second dimension represents the number of risk allele (Alt allele), the value of both dimensions of missing genotype is set to zero. **(c)**, We first groups SNP neurons according to chromosomes, then adopt a partial connected layer to connect SNP feature vectors to corresponding genes and use BiLSTM to capture the interaction relationship between genes. The deep polygenic risk score is obtained based on the fully connected layer with a sigmoid function.

**Table 1.**
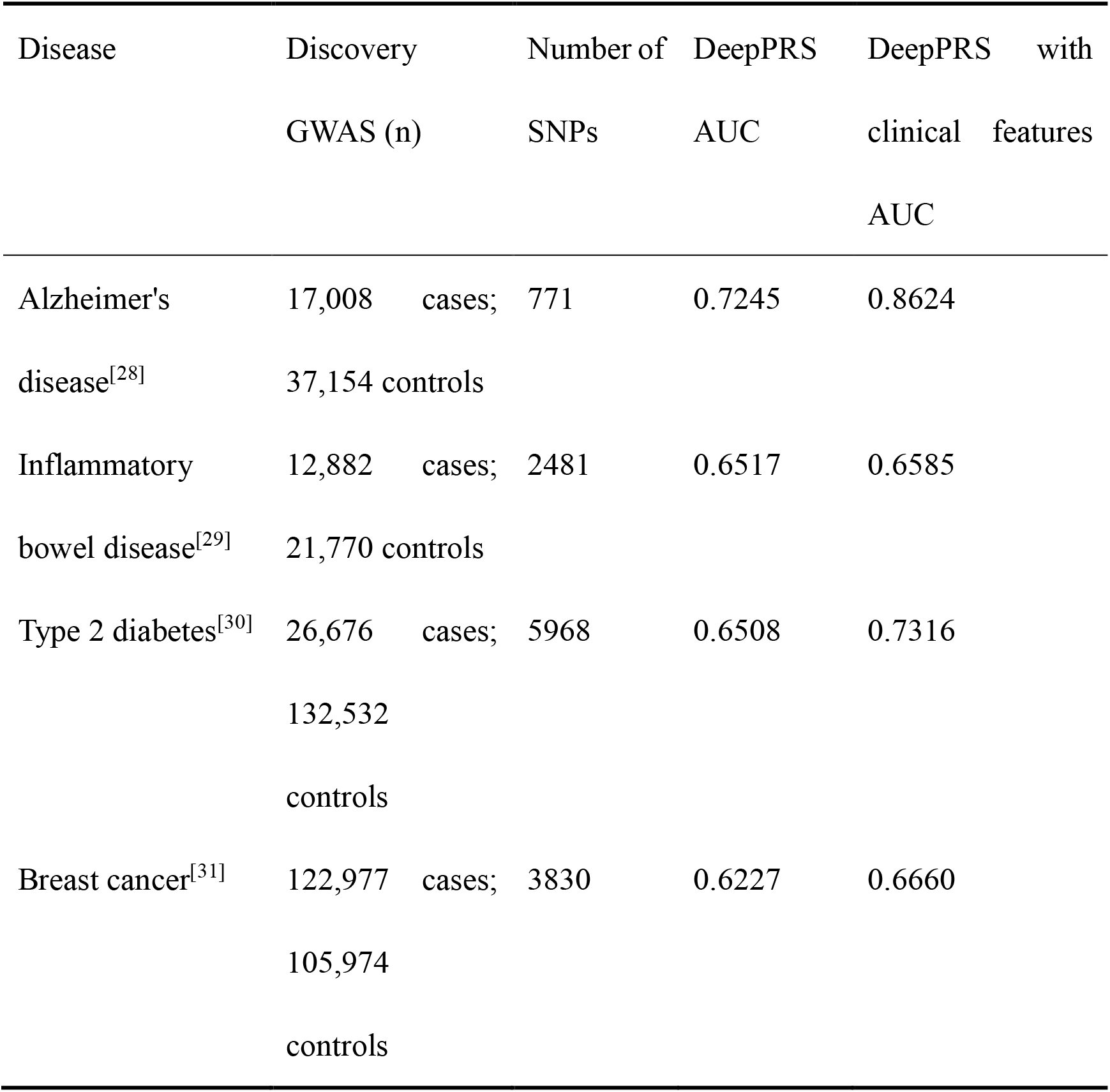
Deep polygenic risk score derivation and testing for four common diseases. Clinical features include age, sex, genotype measurement batch, genotype array, region of assessment center, Townsend Deprivation index at recruitment, education-qualifications, the first four of genetic principal components. The breast cancer analysis is restricted to female participants.

We have studied four common diseases, including Alzheimer’s disease (AD), inflammatory bowel disease (IBD), type 2 diabetes (T2D) and breast cancer (BC). The basic information of the dataset is shown in Table 1. The Area Under the Receiver Operating Characteristic Curve (AUC)^[33]^ of deep polygenic risk score is 0.7245, 0.6517, 0.6508 and 0.6227 for Alzheimer’s disease, inflammatory bowel disease, type 2 diabetes and breast cancer in the test set, respectively (**Table 1**). The deep polygenic risk score with clinical features also performs well in the test dataset, with AUC of 0.8624, 0.6585, 0.7316 and 0.6660 for Alzheimer’s disease, inflammatory bowel disease, type 2 diabetes and breast cancer, respectively (**Table 1**). Details of DeepPRS are provided in the Methods section.

### 2.2. DeepPRS can identify individuals with high risk for AD, IBD, T2D and BC

We perform a series of risk analyses to test whether DeepPRS can identify individuals with high disease risk. Similar to the method used in previous study^[1]^, a given threshold is used to separate the individuals based on deep polygenic risk score. The odds ratio (OR) is used to measure the association between individuals with high PRS versus remainders of population, and the percentile score is used to measure the location of data (see details in the Methods section). Taking IBD as an example, the deep polygenic risk score of IBD is normally distributed across all tested individuals. We find that 4.19% of the population have inherited a genetic predisposition with more than threefold increased risk for IBD. Furthermore, 0.9% of the population have more than fourfold increased risk for IBD and 0.29% have more than fivefold increased risk (**Figure. 2a** and **Table 2**). The median DeepPRSIBD percentile score is 69 for individuals with IBD, which is much higher than the 49 for non-IBD individuals (**Figure. 2b**). The dataset is divided into 100 groups according to the percentile of deep polygenic risk score. The risk of IBD increases sharply in the right tail of the deep polygenic score distribution, from 0.64% in the lowest percentile to 6.24% in the highest percentile, which shows the good risk prediction ability of DeepPRS (**Figure. 2c**). Similar results can also be found on AD, BC and T2D (**Supplementary Figure. S2-4**). The proportion of the population with OR greater than five are 7.90%, 0.06% and 0.15% for AD, BC and T2D, respectively (**Supplementary Figure. S2a-4a**). The median DeepPRS percentile scores are 81, 65, 67 for individuals with AD, BC and T2D respectively, which is much higher than the 49, 47, 48 for non-AD, non-BC and non-T2D individuals (**Supplementary Figure. S2b-4b**). The risks of AD, BC and T2D all increase sharply in the right tail of the deep polygenic score distribution, from 0.17%, 2.85%, 2.31% in the lowest percentile to 4.70%, 23.13%, 26.10% in the highest percentile respectively (**Supplementary Figure. S2c-4c**). Furthermore, DeepPRS performs substantially better than the lasso model for different given thresholds of defining high risk individuals (**Supplementary Table S1**). For top 1% of population versus the others, using the deep polygenic risk score with genotype information only, the ORs of AD, IBD, T2D and BC are 8.97, 3.75, 3.70 and 3.29 respectively. For the lasso model, the ORs are much lower, which are 3.58, 2.15, 0.71 and 1.68 for AD, IBD, T2D and BC respectively.

**Figure. 2.**
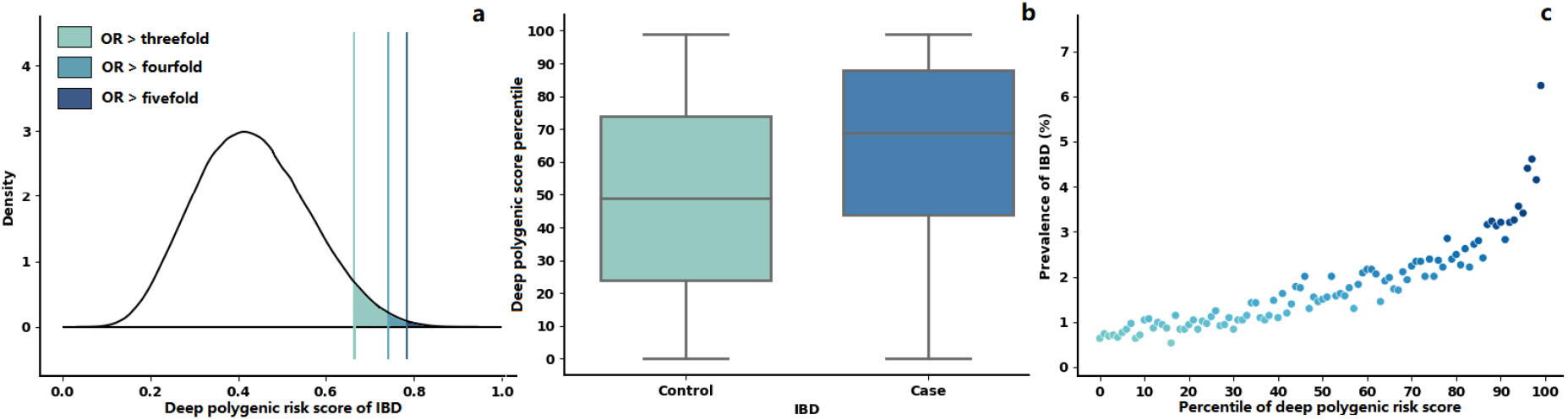
Risk analysis for IBD based on DeepPRS. **(a)**, Distribution of deep polygenic risk score of IBD in the UK Biobank dataset. The x axis represents deep polygenic risk score of IBD. Shading reflects the proportion of the population with three-, four-, and fivefold increased risk versus the remainder of the population. **(b)**, Deep polygenic risk score percentile among IBD cases versus controls in the UK Biobank dataset. In each boxplot, the horizontal lines reflect the median, the top and bottom of each box reflect the quartile range, the whiskers reflect the maximum and minimum values within each group. **(c)**, Prevalence of IBD according to 100 groups of the dataset binned according to the percentile of the deep polygenic risk score of IBD.

**Table 2.** Proportion of the population at three-, four- and fivefold increased risk for Alzheimer’s disease, inflammatory bowel disease, type 2 diabetes and breast cancer.

| High PRS definition | Individuals in UKB dataset (n) | % of individuals |
| --- | --- | --- |
| **Odds ratio $\geq 3.0$** |  |  |
| Alzheimer's disease | 243,567/351,022 | 69.39 |
| Inflammatory bowel disease | 16,457/392,613 | 4.19 |
| Type 2 diabetes | 14,190/346,333 | 4.10 |
| Breast cancer | 2,731/172,160 | 1.59 |
| **Odds ratio $\geq 4.0$** |  |  |
| Alzheimer's disease | 140,151/351,022 | 39.93 |
| Inflammatory bowel disease | 3,515/392,613 | 0.90 |
| Type 2 diabetes | 2,283/346,333 | 0.66 |
| Breast cancer | 328/172,160 | 0.19 |
| **Odds ratio $\geq 5.0$** |  |  |
| Alzheimer's disease | 27,706/351,022 | 7.90 |
| Inflammatory bowel disease | 1,138/392,613 | 0.29 |
| Type 2 diabetes | 532/346,333 | 0.15 |
| Breast cancer | 103/172,160 | 0.06 |

### 2.3. DeepPRS performs better than other two existing methods on four diseases

We study four common diseases, including AD, IBD, T2D and BC on UKB dataset. UKB dataset is divided into case group and control group according to the specific disease (**Table 1** and **Supplementary Figure. S5-8**). To avoid logic circle, we use the GWAS results that do not include UKB dataset. Taking AD as an example, the SNP selection is based on a recent GWAS involving 54,162 participants and a linkage disequilibrium reference panel of 503 Europeans from 1000 Genomes (**Table 1**). The UK Biobank dataset after quality control includes 351,022 participants, of which 2,066 are diagnosed with AD (**Table 1** and **Supplementary Figure. S5**). When *P* < 0.0005 and *r*^*2*^ < 1, the prediction performance is the best, including 771 variants (**Table 1**). On the task of disease risk prediction, we use ten-fold cross-validation^[34]^ for performance comparison (**Table 1** and **Figure. 3**). We predict the disease risk based on genotype only and genotype with clinical features using three methods, including DeepPRS, pruning and thresholding method^[21]^, and lasso model^[22]^ (**Supplementary Table S2-9**). Compared with the other two state-of-art methods, the results show that the DeepPRS achieves the best performance using genotype only or genotype with clinical features as input on all four diseases. The results also show that DeepPRS is not sensitive to the linkage disequilibrium parameters (**Supplementary Table S2-9**), indicating that DeepPRS can automatically deal with the redundancy information between different SNPs without adding more prior information for linkage disequilibrium filtering.

**Figure. 3.**
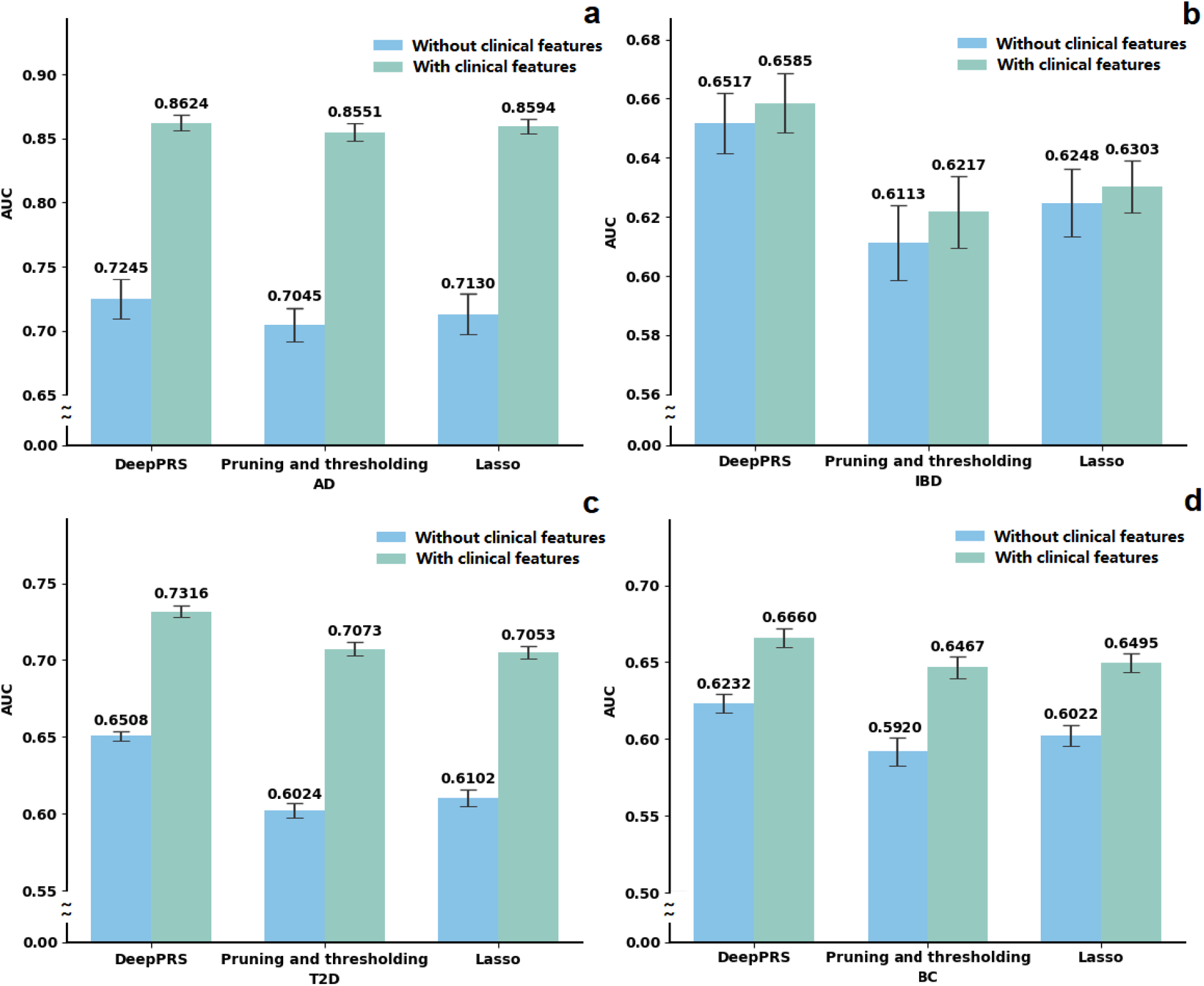
The results of risk prediction using genotype only and genotype with clinical features for Alzheimer’s disease (a), inflammatory bowel disease (b), type 2 diabetes (c) and breast cancer (d). Compared with the pruning and thresholding method, and lasso model, the results show that the DeepPRS achieves the best performance on all the four diseases.

### 2.4. DeepPRS can identify the cases with few known risk SNPs

The traditional weighted-sum methods for calculating PRS are mainly based on the number of risk SNPs and their effect estimate (*β*) from GWAS result. Comparing with previous method, DeepPRS can find cases (diagnosed individuals) with few known risk SNPs. Taking AD as an example, three individuals in the case group only carry few risk SNPs. Not surprisingly, their PRS percentile scores based on pruning and thresholding method are very low, which are all 1. However, their DeepPRS percentile scores are all 99, which are larger than threshold of the 9.53-fold risk (**Supplementary Table S10**). For IBD, PRS percentile scores based on pruning and thresholding method of three individuals are 5, 5 and 6, respectively. However, their DeepPRS percentile scores are all 99, which are larger than threshold of the 4.23-fold risk (**Supplementary Table S10**). For T2D, PRS percentile scores based on pruning and thresholding method of three individuals are 1, 2 and 9, respectively. However, their DeepPRS percentile scores are 75, 98 and 86, which are larger than threshold of the 2.34-fold risk (**Supplementary Table S10**). For BC, PRS percentile scores based on pruning and thresholding method of three individuals are 5, 19 and 9, respectively. However, their DeepPRS percentile scores are 75, 98 and 86, which are larger than threshold of the 2.23-fold risk (**Supplementary Table S10**).

### 2.5. Each component of DeepPRS is well-designed for diseases risk prediction

In order to illustrate the contribution of each part of DeepPRS to the prediction performance, we test three varied methods by replacing three parts of the DeepPRS respectively, including using the traditional additive encoding method to replace our encoding schema (method a), directly using SNP features for subsequent feature extraction without the partial connected layer (method b), using CNN to replace BiLSTM to extract gene features (method c). In this test, we only use genotype features as input, and compare different methods based on ten-fold cross validation. The results show that the performance of DeepPRS is better than the other three methods, indicating that the model is designed properly (**Table 3**). Compared with the additive encoding schema of method a, the reason for the superior performance of DeepPRS may be that the new encoding method expands the dimension of the feature. The traditional encoding method uses 0, 1 and 2 to represent genotypes, which is likely to bring additional quantitative relationships. In fact, there may not be a two-fold risk relationship between homozygous genotype and heterozygous genotype. We remove this additional quantitative relationship and better handle the missing genotype by filling it with (zero, zero) instead of deleting the locus. Compared with method b, DeepPRS uses partial connected layer to connect the SNP features to the neuron representing their adjacent gene. It not only greatly reduces the number of parameters and running time of deep neural networks model, but also takes the association between SNPs and genes into account. In addition, the effect of SNPs on the individuals is ultimately expressed by genes, and it is of more biological significance to extract features using gene-based partial connected layer. Compared with the CNN model of method c, the reason for the superior performance of DeepPRS may due to the fact that CNN can only extract local SNPs interaction relations, while BiLSTM can capture SNP interaction information at a distance on a chromosome.

**Table 3.** The effect of each component of DeepPRS on disease risk prediction.

|  | DeepPRS | Method a | Method b | Method c |
| --- | --- | --- | --- | --- |
| Disease | AUC | AUC | AUC | AUC |
| **Alzheimer's disease** | 0.7245 | 0.7158 | 0.7216 | 0.7232 |
| **Inflammatory bowel disease** | 0.6517 | 0.6485 | 0.6469 | 0.6473 |
| **Type 2 diabetes** | 0.6508 | 0.6490 | 0.6440 | 0.6457 |
| **Breast cancer** | 0.6227 | 0.6169 | 0.6232 | 0.6162 |

### 2.6. DeepPRS is robust for the variation of significant SNPs on disease risk prediction

At present, GWAS has not identified all significant loci. However, DeepPRS, pruning and thresholding method and Lasso method all need to select significant SNPs based on the *P* value given by the GWAS. Therefore, we test whether the absence of significant SNPs may affect the performance of these three methods. We sequentially removed the most significant SNPs in the top 10, top 20 and top 50 from GWAS result. The results show that DeepPRS performs consistently better than the other two methods (**Figure. 4**). Although the AUCs of all three methods decrease with the increase of missing significant SNPs, DeepPRS is the most robust one (**Figure. 4**). For example, the decrease of AUC for DeepPRS is more than two times slower than pruning and thresholding method on breast cancer (**Figure. 4d**). In summary, compared with the pruning and thresholding method, and lasso method, the results show that the DeepPRS is more robust for the variation of significant SNPs on disease risk prediction.

**Figure. 4.**
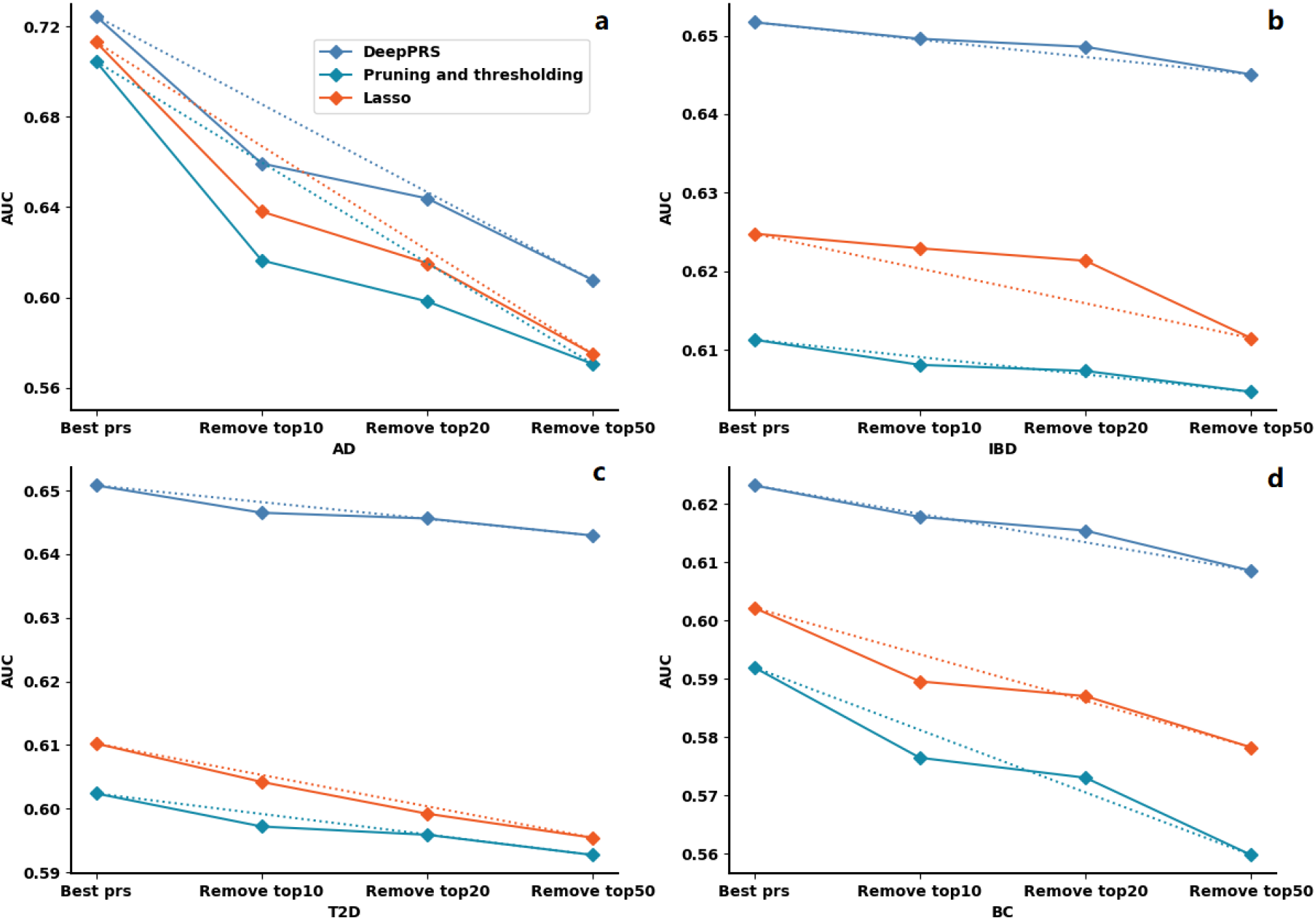
The results of risk prediction for Alzheimer’s disease(a), inflammatory bowel disease (b), type 2 diabetes (c) and breast cancer (d) after removing different significant SNPs. The dotted line indicates the rate of performance degradation. The *X* axis represents the number of significant SNPs removed in different evaluation tests, and the *Y* axis represents the AUC score.

## 3. Discussion

In the past 25 years, the clinical potential of PRS has attracted widespread attention. The success of PRS depends on not only the genome-wide association study (GWAS) data but also the development of risk models. More accurate disease risk prediction may lead to better customize screening, prevention and treatment. In this study, we propose a novel PRS calculation model and analyze the risk of four common diseases. Alzheimer’s disease, as the most common form of dementia, accounts for about 60% of cases of dementia^[35]^. At present, there are about 50 million AD patients worldwide, which has become one of the biggest public health challenges. Early diagnosis of AD may bring important personal and economic benefits^[36]^. DeepPRSAD identifies 7.90% of the population at greater than fivefold risk and the top 1% of the population have more than 8.97-fold risk (**Supplementary Table S1**). For AD, the proportion of individuals at high risk is much higher than the other three diseases, which may be due to the fact that the number of cases in the AD data set is too small. As a risk warning, this score can be a sign of early intervention for high-risk people. The results of a large, long-term, randomized controlled trial show that multifaceted interventions, including diet, exercise, cognitive training, and vascular risk monitoring, can improve or maintain cognitive function in the general dementia risk population^[37]^ (60-77 years old).

As a global disease with accelerating incidence in newly industrialized countries, the burden of IBD remains high and prevalence surpasses 0.3%^[38][39]^. DeepPRSIBD identifies 4.19% of the population at greater than threefold risk and the top 1% have more than 3.75-fold risk (**Supplementary Table S1**). Although existing studies have shown that factors such as diet, probiotics, and antibiotics are related to the development of IBD, more studies are still needed to explore the mechanisms that can help to prevent IBD^[38][40]^. The high-risk population identify by DeepPRS may bring new opportunities for large-scale population epidemiological studies to assess a novel preventive therapy.

Type 2 diabetes is an expanding global health problem and puts a huge burden on health-care systems^[41]^. DeepPRST2D identifies 4.10% of the population at greater than threefold risk and the top 1% have more than 3.70-fold risk (**Supplementary Table S1**). A study from Finland found that increasing physical activity and an intensive lifestyle may substantially reduce the incidence of type 2 diabetes in high-risk individuals^[42]^. Therefore, the high-risk population determined by our DeepPRST2D can take early prevention to reduce the risk of type 2 diabetes.

Breast cancer is the second leading cause of death from cancer in women, affecting one in twenty globally and as many as one in eight in high-income countries^[43]^. Fortunately, studies have shown that early detection and treatment can considerably improve outcomes of patients^[43][44]^. DeepPRSBC identifies 1.59% of the population at greater than threefold risk and the top 1% have more than 3.28-fold risk (**Supplementary Table S1**). Although the current prevention methods continue to increase, the overdiagnosis of screening, the serious side effects of chemical and biological prevention still cannot be ignored^[45]^. Ascertainment of those with high deep polygenic risk score may provide an opportunity to adopt these interventions more precisely.

These results show that the DeepPRS can identify higher-risk individuals in the population for several common diseases. In addition to using the genotype information available at birth as a predictor, more clinical features can be added to the predictor over time. Then, the accuracy of the DeepPRS may be improved.

## Data Availability

All data produced in the present study are available upon reasonable request to the authors.

## Acknowledgments

This research is conducted with approved access to UK Biobank data under application number 53464.

## Funding

National Natural Science Foundation of China grant No. 62072376 and U1811262

## Author contributions

Conceptualization: JJ.P, Z.W, X.S

Methodology: JJ.P, J.L, R.H, Z.W

Investigation: J.L, R.H, Y.W, L.H, JH.P

Supervision: JJ.P, T.W, J.H, X.S, Z.W

Writing—original draft: JJ.P, J.L, R.H, Y.W

Writing—review & editing: JJ.P, J.L, T.W, J.H, X.S, Z.W

## Competing interests

Authors declare that they have no competing interests.

## Data and materials availability

All data used in the analyses are available in UK Biobank at https://www.ukbiobank.ac.uk/. The code will be publicly released after the paper is accepted.

## Notes

### Competing Interest Statement

The authors have declared no competing interest.

### Funding Statement

This study was funded by National Natural Science Foundation of China grant No. 62072376 and U1811262.

