## Supplementary Document for "A Deep Learning-based Genome-wide Polygenic Risk Score for Common Diseases Identifies Individuals with Risk"

### DeepPRS model with clinical features

On the basis of the DeepPRS model, SNP features are firstly transformed into gene features by the partial connected layer. The number of clinical features is much smaller than the number of genes. The contribution of clinical features in the neural network will be limited and may have little effect on the final prediction performance, if we concatenate the clinical features with gene features directly. Therefore, the normalized covariate features are first expanded to the same dimension as the gene features by the fully connected layer. Then, the gene features and clinical features are concatenated together, and the BiLSTM layer is used for feature extraction. Finally, we concatenate the output of the BiLSTM layer and the low-dimensional clinical featuresbefore the fully connected layer to get the final deep polygenic risk score with clinical features (Figure. S1).

### Parameters of DeepPRS model

The parameters of the BiLSTM are as follows. We set the hidden units of each unidirectional LSTM as 4. In order to avoid overfitting, we add 0.001 of L2 regularization and 0.25 of dropout rate on BiLSTM layer. We run 20 epochs with 512 samples for each batch. We use AdamW algorithm and set an initial learning rate as 0.001 to optimize the weighted binary cross-entropy loss.

### Introduction of two compare methods

### Pruning and thresholding

The pruning and thresholding method calculates polygenic risk score based on the weights and SNP genotypes. Weights are generally assigned to each genetic variant according to the strength of their association with disease risk (effect estimate) from recent GWAS. The genotypes are scored based on how many risk alleles they have for each variant (for example, zero, one, or two). Given an individual having $n$ SNPs, the PRS can be obtained as follows:

$$PRS = \sum_{i=1}^{n} \beta_i x_i \qquad (1)$$

Where the $\beta_i$ is the effect estimate of SNP $i$, $x_i$ is the genotype of SNP $i$. The PRS of the pruning and thresholding method is evaluated based on a logistic regression model with the disease as the outcome. The loss function is the binary cross-entropy loss with class weights.

**Lasso method**

The lasso method calculates PRS based on SNP genotypes. The PRS of the lasso method is obtained based on a LASSO logistic regression model. The loss function is the binary cross-entropy loss with class weights. We used AdamW algorithm and set an initial learning rate as 0.001 to optimize the weighted binary cross-entropy loss.

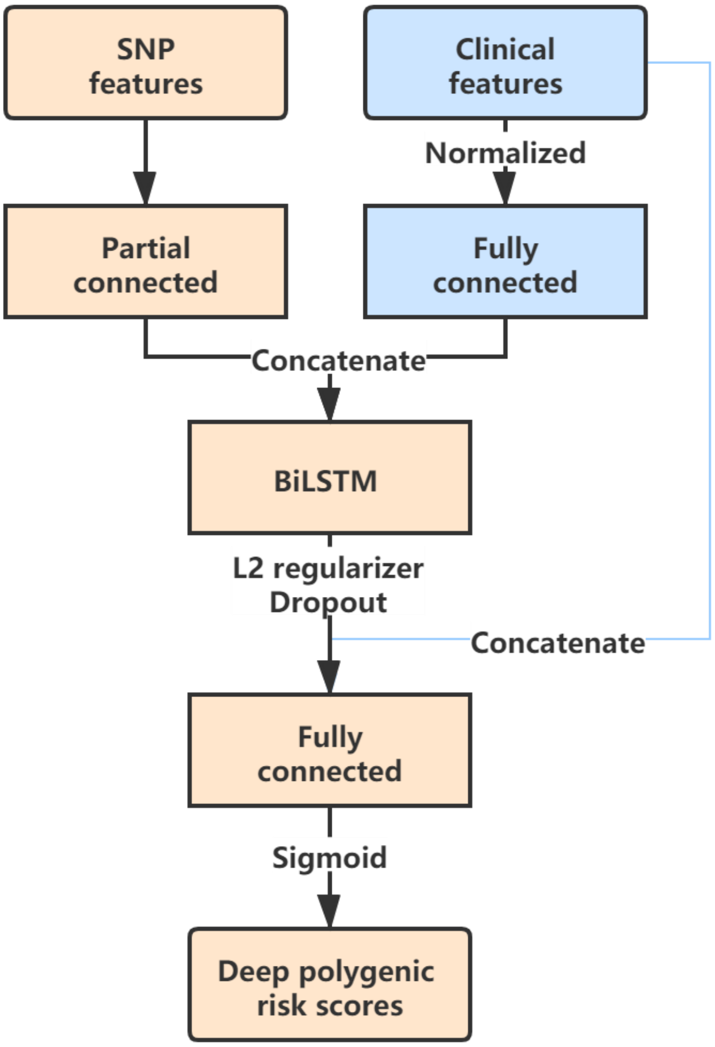


**Figure. S1. The model architecture of DeepPRS with clinical features.**

The deep polygenic risk score with clinical features is determined using the DeepPRS model with genotype, age, sex, genotype measurement batch, genotype array, region of assessment center, Townsend Deprivation index at recruitment, education-qualifications, the first four of genetic principal components. The blue components in this figure are used to incorporate the clinical features with genotype information.

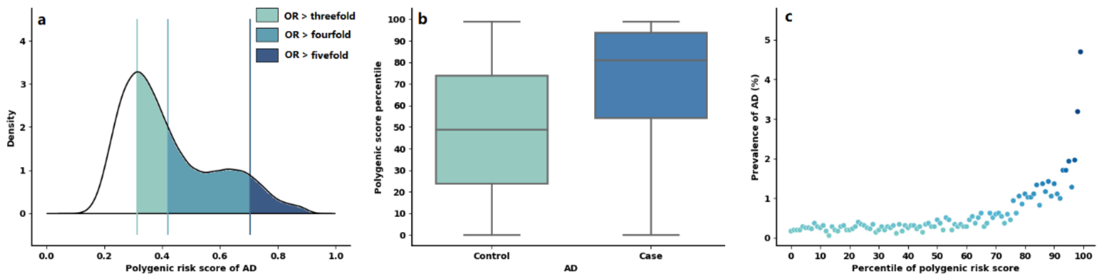


**Figure. S2. Risk analysis for AD based on DeepPRS.**

**(a)**, Distribution of deep polygenic risk score of AD in the UK Biobank dataset. The *X* axis represents deep polygenic risk score of AD. Shading reflects the proportion of the population with three-, four-, and fivefold increased risk versus the remainder of the population. **(b)**, Deep polygenic risk score percentile among AD cases versus controls in the UK Biobank dataset. In each boxplot, the horizontal lines reflect the median. The top and bottom of each box reflect the quartile range. The whiskers reflect the maximum and minimum values within each group. **(c)**, Prevalence of AD according to 100 groups of the dataset binned according to the percentile of the deep polygenic risk score of AD.

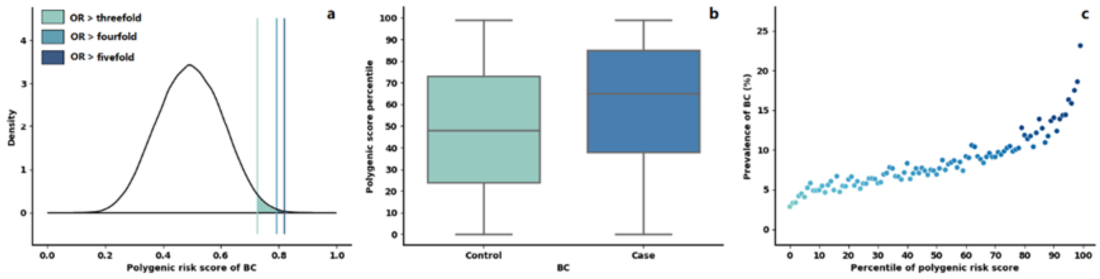


**Figure. S3. Risk analysis for BC based on DeepPRS.**

**(a)**, Distribution of deep polygenic risk score of BC in the UK Biobank dataset. The x axis represents deep polygenic risk score of BC. Shading reflects the proportion of the population with three-, four-, and fivefold increased risk versus the remainder of the population. **(b)**, Deep polygenic risk score percentile among BC cases versus controls in the UK Biobank dataset. In each boxplot, the horizontal lines reflect the median. The top and bottom of each box reflect the quartile range. The whiskers reflect the maximum and minimum values within each group. **(c)**, Prevalence of BC according to 100 groups of the dataset binned according to the percentile of the deep polygenic risk score of BC.

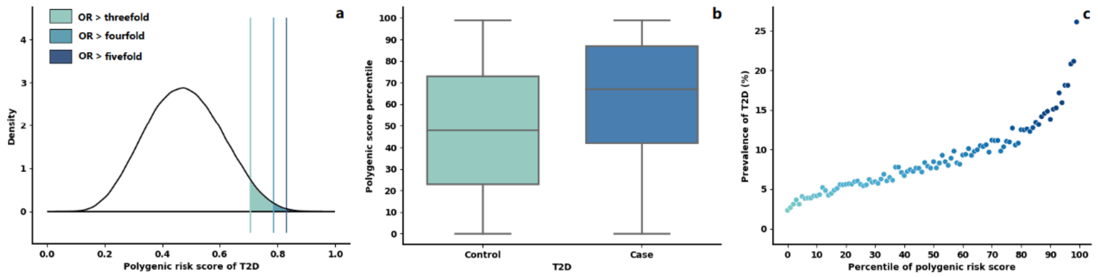


**Figure. S4. Risk analysis for T2D based on DeepPRS.**

**(a)**, Distribution of deep polygenic risk score of T2D in the UK Biobank dataset. The x axis represents deep polygenic risk score of T2D. Shading reflects the proportion of the population with three-, four-, and fivefold increased risk versus the remainder of the population. **(b)**, Deep polygenic risk score percentile among T2D cases versus controls in the UK Biobank dataset. In each boxplot, the horizontal lines reflect the median. The top and bottom of each box reflect the quartile range. The whiskers reflect the maximum and minimum values within each group. **(c)**, Prevalence of T2D according to 100 groups of the dataset binned according to the percentile of the deep polygenic risk score of T2D.

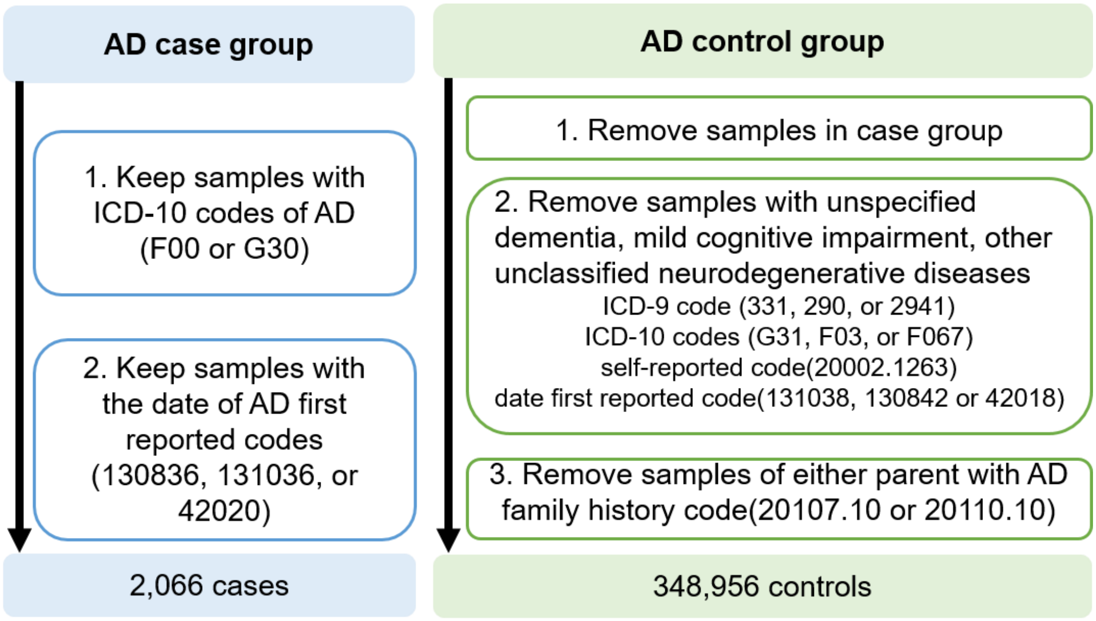


**Figure. S5. Determination of case group and control group for Alzheimer's disease.**

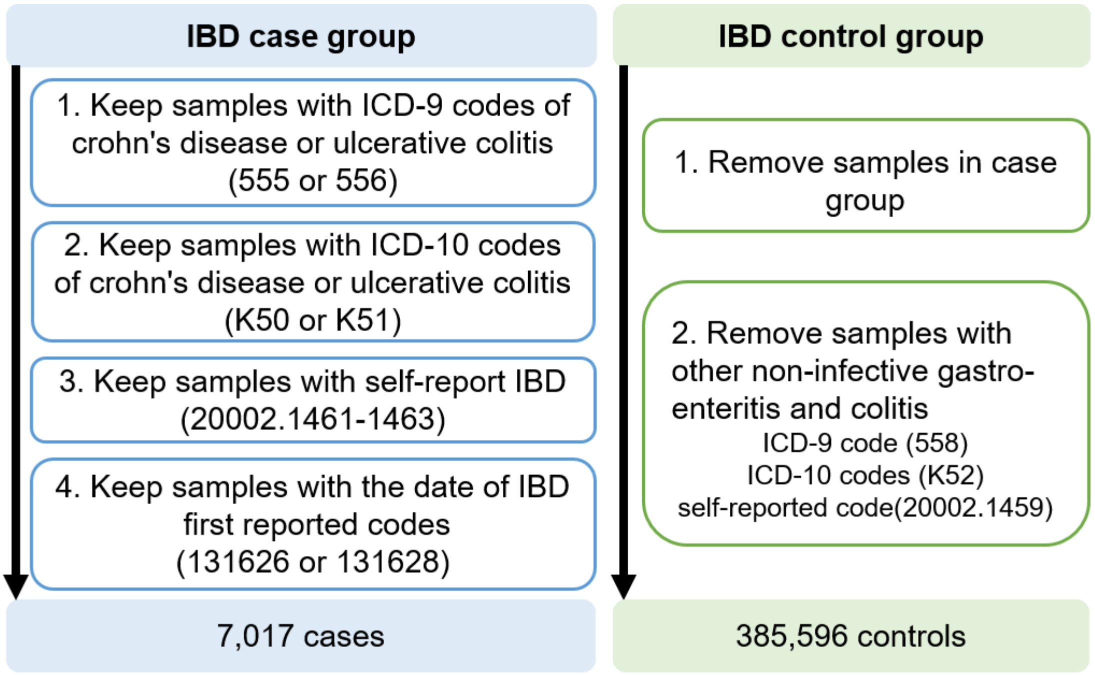


**Figure. S6. Determination of case group and control group for inflammatory bowel disease.**

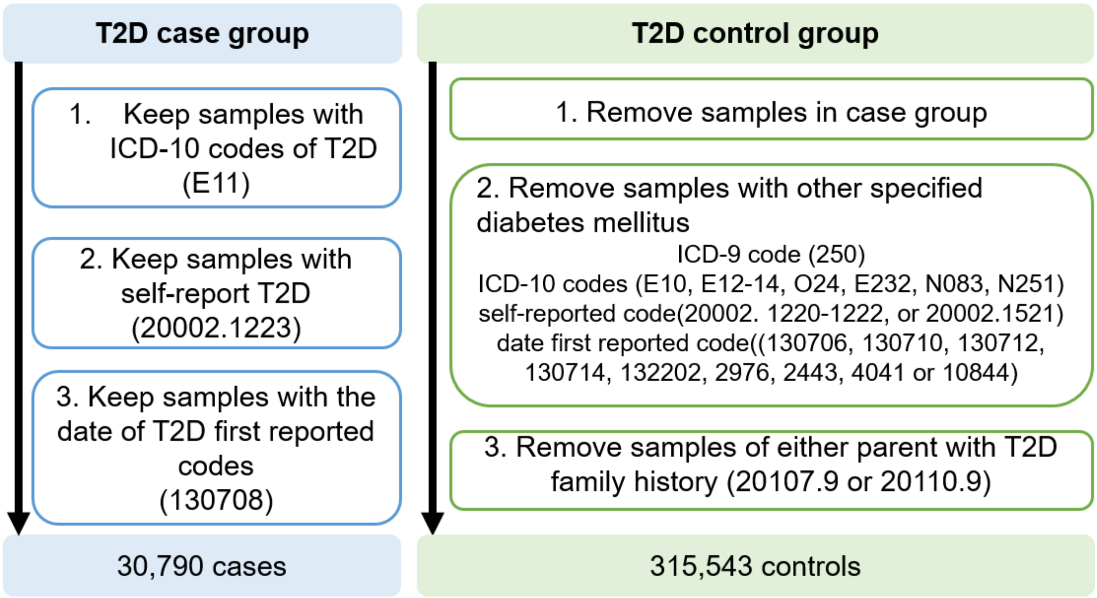


**Figure. S7. Determination of case group and control group for type 2 diabetes.**

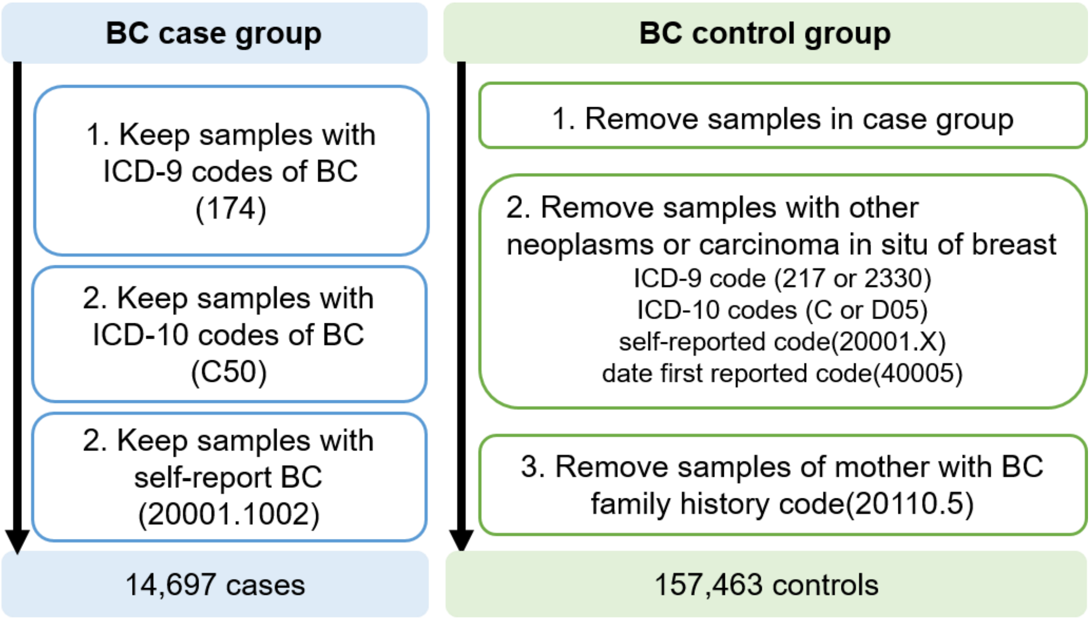


**Figure. S8. Determination of case group and control group for breast cancer.**

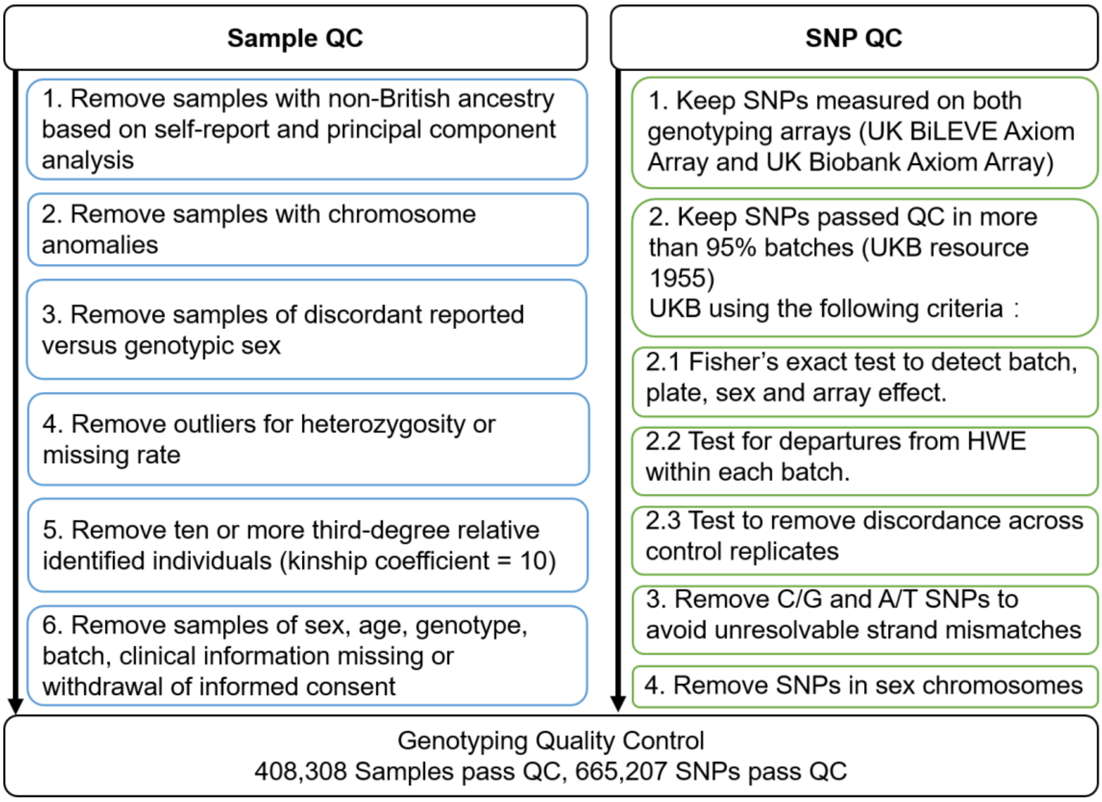


**Figure. S9. Quality control process of the UK Biobank genotype dataset.** The left side includes the sample QC steps. The right side includes the SNP QC steps.

**Table S1. Odds ratio of a high deep polygenic risk score.** The polygenic risk score is generated based on the genotype information in the UKB dataset. The calculation of odds ratio (OR) is based on the polygenetic risk score generated under the best performance parameters of each method.

| High deep polygenic risk score definition | Reference group | OR based on DeepPRS | OR based on lasso model |
|---|---|---|---|
| **Alzheimer's disease** | | | |
| Top 20% of distribution | Remaining 80% | 4.64 | 2.15 |
| Top 10% of distribution | Remaining 90% | 4.70 | 2.45 |
| Top 5% of distribution | Remaining 95% | 5.55 | 2.81 |
| Top 1% of distribution | Remaining 99% | 8.97 | 3.58 |
| **Inflammatory bowel disease** | | | |
| Top 20% of distribution | Remaining 80% | 2.39 | 1.58 |
| Top 10% of distribution | Remaining 90% | 2.56 | 1.78 |
| Top 5% of distribution | Remaining 95% | 2.87 | 1.93 |
| Top 1% of distribution | Remaining 99% | 3.75 | 2.15 |
| **Type 2 diabetes** | | | |
| Top 20% of distribution | Remaining 80% | 2.41 | 0.57 |
| Top 10% of distribution | Remaining 90% | 2.60 | 0.62 |
| Top 5% of distribution | Remaining 95% | 2.92 | 0.67 |
| Top 1% of distribution | Remaining 99% | 3.70 | 0.71 |
| **Breast cancer** | | | |
| Top 20% of distribution | Remaining 80% | 2.12 | 1.22 |
| Top 10% of distribution | Remaining 90% | 2.29 | 1.30 |
| Top 5% of distribution | Remaining 95% | 2.56 | 1.43 |
| Top 1% of distribution | Remaining 99% | 3.29 | 1.68 |

**Table S2. The comparison of risk prediction performance for Alzheimer's disease using only genotype information.** Bold numbers indicate the best performance.

| Tuning Parameter | N Variants in Score | N Genes in Score | DeepPRS AUC | Pruning and thresholding AUC | Lasso AUC |
|---|---|---|---|---|---|
| $P<5\times10^{-3}$; - | 3834 | 1454 | 0.7145 | 0.6354 | 0.7005 |
| $P<5\times10^{-3}$; $r^2<0.8$ | 3291 | 1448 | 0.7170 | 0.6681 | 0.7037 |
| $P<5\times10^{-3}$; $r^2<0.6$ | 3048 | 1444 | 0.7146 | 0.6585 | 0.7029 |
| $P<5\times10^{-3}$; $r^2<0.4$ | 2811 | 1439 | 0.7144 | 0.6440 | 0.7002 |
| $P<5\times10^{-3}$; $r^2<0.2$ | 2577 | 1432 | 0.6995 | 0.6023 | 0.6831 |
| $P<5\times10^{-4}$; - | 771 | 291 | **0.7245** | 0.6765 | 0.7108 |
| $P<5\times10^{-4}$; $r^2<0.8$ | 606 | 287 | 0.7237 | 0.6983 | **0.7130** |
| $P<5\times10^{-4}$; $r^2<0.6$ | 550 | 287 | 0.7212 | 0.6941 | 0.7105 |
| $P<5\times10^{-4}$; $r^2<0.4$ | 491 | 287 | 0.7186 | 0.6882 | 0.7098 |
| $P<5\times10^{-4}$; $r^2<0.2$ | 440 | 288 | 0.7035 | 0.6544 | 0.6929 |
| $P<5\times10^{-5}$; - | 259 | 57 | 0.7212 | 0.6972 | 0.7100 |
| $P<5\times10^{-5}$; $r^2<0.8$ | 194 | 57 | 0.7205 | 0.7039 | 0.7095 |
| $P<5\times10^{-5}$; $r^2<0.6$ | 169 | 57 | 0.7187 | 0.7016 | 0.7078 |
| $P<5\times10^{-5}$; $r^2<0.4$ | 146 | 58 | 0.7169 | 0.6998 | 0.7086 |
| $P<5\times10^{-5}$; $r^2<0.2$ | 115 | 57 | 0.7029 | 0.6747 | 0.6924 |
| $P<5\times10^{-6}$; - | 131 | 25 | 0.7180 | **0.7045** | 0.7093 |
| $P<5\times10^{-6}$; $r^2<0.8$ | 105 | 25 | 0.7173 | 0.7034 | 0.7089 |
| $P<5\times10^{-6}$; $r^2<0.6$ | 88 | 25 | 0.7154 | 0.7014 | 0.7074 |
| $P<5\times10^{-6}$; $r^2<0.4$ | 77 | 25 | 0.7134 | 0.6996 | 0.7079 |

| | | | | | |
|---|---|---|---|---|---|
| $P<5\times10^{-6}$; $r^2<0.2$ | 62 | 24 | 0.7010 | 0.6767 | 0.6924 |

**Table S3. The comparison of risk prediction performance for Alzheimer's disease using genotype information and clinical features.** Bold numbers indicate the best performance.

| Tuning Parameter | N Variants in Score | N Genes in Score | DeepPRS with clinical features AUC | Pruning and thresholding with clinical features AUC | Lasso with clinical features AUC |
|---|---|---|---|---|---|
| $P<5\times10^{-3}$; - | 3834 | 1454 | 0.8586 | 0.8322 | 0.8477 |
| $P<5\times10^{-3}$; $r^2<0.8$ | 3291 | 1448 | 0.8581 | 0.8419 | 0.8514 |
| $P<5\times10^{-3}$; $r^2<0.6$ | 3048 | 1444 | 0.8591 | 0.8387 | 0.8523 |
| $P<5\times10^{-3}$; $r^2<0.4$ | 2811 | 1439 | 0.8588 | 0.8344 | 0.8540 |
| $P<5\times10^{-3}$; $r^2<0.2$ | 2577 | 1432 | 0.8525 | 0.8237 | 0.8471 |
| $P<5\times10^{-4}$; - | 771 | 291 | **0.8624** | 0.8444 | 0.8591 |
| $P<5\times10^{-4}$; $r^2<0.8$ | 606 | 287 | 0.8621 | 0.8519 | 0.8592 |
| $P<5\times10^{-4}$; $r^2<0.6$ | 550 | 287 | 0.8615 | 0.8504 | 0.8592 |
| $P<5\times10^{-4}$; $r^2<0.4$ | 491 | 287 | 0.8603 | 0.8484 | 0.8586 |
| $P<5\times10^{-4}$; $r^2<0.2$ | 440 | 288 | 0.8538 | 0.8370 | 0.8508 |
| $P<5\times10^{-5}$; - | 259 | 57 | 0.8614 | 0.8517 | 0.8594 |
| $P<5\times10^{-5}$; $r^2<0.8$ | 194 | 57 | 0.8615 | 0.8546 | 0.8592 |
| $P<5\times10^{-5}$; $r^2<0.6$ | 169 | 57 | 0.8612 | 0.8537 | 0.8588 |
| $P<5\times10^{-5}$; $r^2<0.4$ | 146 | 58 | 0.8605 | 0.8529 | 0.8581 |
| $P<5\times10^{-5}$; $r^2<0.2$ | 115 | 57 | 0.8537 | 0.8439 | 0.8508 |
| $P<5\times10^{-6}$; - | 131 | 25 | 0.8608 | **0.8551** | 0.8593 |
| $P<5\times10^{-6}$; $r^2<0.8$ | 105 | 25 | 0.8610 | 0.8548 | **0.8594** |
| $P<5\times10^{-6}$; $r^2<0.6$ | 88 | 25 | 0.8605 | 0.8540 | 0.8587 |

| | | | | | |
|---|---|---|---|---|---|
| $P<5\times10^{-6}$; $r^2<0.4$ | 77 | 25 | 0.8595 | 0.8531 | 0.8584 |
| $P<5\times10^{-6}$; $r^2<0.2$ | 62 | 24 | 0.8536 | 0.8448 | 0.8509 |

**Table S4. The comparison of risk prediction performance for inflammatory bowel disease using only genotype information.** Bold numbers indicate the best performance.

| Tuning Parameter | N Variants in Score | N Genes in Score | DeepPRS AUC | Pruning and thresholding AUC | Lasso AUC |
|---|---|---|---|---|---|
| $P<5\times10^{-3}$; - | 7848 | 2082 | 0.6506 | 0.5817 | 0.6157 |
| $P<5\times10^{-3}$; $r^2<0.8$ | 6188 | 2070 | 0.6503 | 0.6005 | 0.6235 |
| $P<5\times10^{-3}$; $r^2<0.6$ | 5626 | 2065 | 0.6501 | 0.6040 | 0.6244 |
| $P<5\times10^{-3}$; $r^2<0.4$ | 5041 | 2055 | 0.6478 | 0.6031 | 0.6211 |
| $P<5\times10^{-3}$; $r^2<0.2$ | 4423 | 2035 | 0.6417 | 0.5962 | 0.6172 |
| $P<5\times10^{-4}$; - | 2481 | 542 | **0.6517** | 0.5783 | 0.6166 |
| $P<5\times10^{-4}$; $r^2<0.8$ | 1689 | 533 | 0.6493 | 0.5986 | 0.6242 |
| $P<5\times10^{-4}$; $r^2<0.6$ | 1438 | 537 | 0.6490 | 0.6062 | **0.6248** |
| $P<5\times10^{-4}$; $r^2<0.4$ | 1219 | 535 | 0.6477 | **0.6113** | 0.6241 |
| $P<5\times10^{-4}$; $r^2<0.2$ | 1018 | 525 | 0.6420 | 0.6099 | 0.6200 |
| $P<5\times10^{-5}$; - | 1298 | 200 | 0.6477 | 0.5750 | 0.6192 |
| $P<5\times10^{-5}$; $r^2<0.8$ | 813 | 198 | 0.6469 | 0.5919 | 0.6201 |
| $P<5\times10^{-5}$; $r^2<0.6$ | 649 | 198 | 0.6477 | 0.6009 | 0.6226 |
| $P<5\times10^{-5}$; $r^2<0.4$ | 535 | 197 | 0.6451 | 0.6087 | 0.6217 |
| $P<5\times10^{-5}$; $r^2<0.2$ | 407 | 193 | 0.6410 | 0.6101 | 0.6184 |
| $P<5\times10^{-6}$; - | 892 | 112 | 0.6410 | 0.5728 | 0.6109 |
| $P<5\times10^{-6}$; $r^2<0.8$ | 535 | 113 | 0.6406 | 0.5874 | 0.6176 |
| $P<5\times10^{-6}$; $r^2<0.6$ | 418 | 113 | 0.6404 | 0.5957 | 0.6185 |
| $P<5\times10^{-6}$; $r^2<0.4$ | 328 | 115 | 0.6380 | 0.6042 | 0.6153 |

| | | | | | |
|---|---|---|---|---|---|
| *P*<5×$10^{-6}$; $r^2$<0.2 | 250 | 113 | 0.6344 | 0.6062 | 0.6136 |

**Table S5. The comparison of risk prediction performance for inflammatory bowel disease using genotype information and clinical features.** Bold numbers indicate the best performance.

| Tuning Parameter | N Variants in Score | N Genes in Score | DeepPRS with clinical features AUC | Pruning and thresholding with clinical features AUC | Lasso with clinical features AUC |
|---|---|---|---|---|---|
| $P<5\times10^{-3}$; - | 7848 | 2082 | 0.6564 | 0.5959 | 0.5772 |
| $P<5\times10^{-3}$; $r^2<0.8$ | 6188 | 2070 | 0.6567 | 0.6125 | 0.6186 |
| $P<5\times10^{-3}$; $r^2<0.6$ | 5626 | 2065 | 0.6568 | 0.6156 | 0.5953 |
| $P<5\times10^{-3}$; $r^2<0.4$ | 5041 | 2055 | 0.6551 | 0.6146 | 0.6114 |
| $P<5\times10^{-3}$; $r^2<0.2$ | 4423 | 2035 | 0.6491 | 0.6083 | 0.6236 |
| $P<5\times10^{-4}$; - | 2481 | 542 | **0.6585** | 0.5929 | 0.6148 |
| $P<5\times10^{-4}$; $r^2<0.8$ | 1689 | 533 | 0.6564 | 0.6104 | 0.6269 |
| $P<5\times10^{-4}$; $r^2<0.6$ | 1438 | 537 | 0.6553 | 0.6172 | 0.6299 |
| $P<5\times10^{-4}$; $r^2<0.4$ | 1219 | 535 | 0.6544 | **0.6217** | 0.6253 |
| $P<5\times10^{-4}$; $r^2<0.2$ | 1018 | 525 | 0.6495 | 0.6206 | 0.6160 |
| $P<5\times10^{-5}$; - | 1298 | 200 | 0.6547 | 0.5897 | 0.6153 |
| $P<5\times10^{-5}$; $r^2<0.8$ | 813 | 198 | 0.6549 | 0.6042 | 0.6211 |
| $P<5\times10^{-5}$; $r^2<0.6$ | 649 | 198 | 0.6552 | 0.6120 | **0.6303** |
| $P<5\times10^{-5}$; $r^2<0.4$ | 535 | 197 | 0.6521 | 0.6191 | 0.6293 |
| $P<5\times10^{-5}$; $r^2<0.2$ | 407 | 193 | 0.6492 | 0.6206 | 0.6283 |
| $P<5\times10^{-6}$; - | 892 | 112 | 0.6485 | 0.5880 | 0.6227 |
| $P<5\times10^{-6}$; $r^2<0.8$ | 535 | 113 | 0.6483 | 0.6004 | 0.6174 |
| $P<5\times10^{-6}$; $r^2<0.6$ | 418 | 113 | 0.6483 | 0.6074 | 0.6228 |

| | | | | | |
|---|---|---|---|---|---|
| $P<5\times10^{-6}$; $r^2<0.4$ | 328 | 115 | 0.6459 | 0.6149 | 0.6267 |
| $P<5\times10^{-6}$; $r^2<0.2$ | 250 | 113 | 0.6427 | 0.6167 | 0.6238 |

**Table S6. The comparison of risk prediction performance for type 2 diabetes using only genotype information.** Bold numbers indicate the best performance**.**

| Tuning Parameter | N Variants in Score | N Genes in Score | DeepPRS AUC | Pruning and thresholding AUC | Lasso AUC |
|---|---|---|---|---|---|
| $P<5\times10^{-3}$; - | 5968 | 1890 | **0.6508** | 0.5899 | **0.6102** |
| $P<5\times10^{-3}$; $r^2<0.8$ | 5069 | 1878 | 0.6501 | **0.6024** | 0.6087 |
| $P<5\times10^{-3}$; $r^2<0.6$ | 4588 | 1865 | 0.6488 | 0.5979 | 0.6080 |
| $P<5\times10^{-3}$; $r^2<0.4$ | 4203 | 1855 | 0.6476 | 0.5958 | 0.6071 |
| $P<5\times10^{-3}$; $r^2<0.2$ | 3769 | 1837 | 0.6440 | 0.5863 | 0.6032 |
| $P<5\times10^{-4}$; - | 1384 | 481 | 0.6346 | 0.5885 | 0.6039 |
| $P<5\times10^{-4}$; $r^2<0.8$ | 1059 | 478 | 0.6338 | 0.6023 | 0.6029 |
| $P<5\times10^{-4}$; $r^2<0.6$ | 919 | 477 | 0.6335 | 0.6006 | 0.6033 |
| $P<5\times10^{-4}$; $r^2<0.4$ | 816 | 475 | 0.6321 | 0.5981 | 0.6012 |
| $P<5\times10^{-4}$; $r^2<0.2$ | 700 | 472 | 0.6300 | 0.5921 | 0.5993 |
| $P<5\times10^{-5}$; - | 538 | 144 | 0.6244 | 0.5950 | 0.5991 |
| $P<5\times10^{-5}$; $r^2<0.8$ | 380 | 144 | 0.6243 | 0.6013 | 0.5984 |
| $P<5\times10^{-5}$; $r^2<0.6$ | 309 | 143 | 0.6232 | 0.6017 | 0.5992 |
| $P<5\times10^{-5}$; $r^2<0.4$ | 263 | 143 | 0.6226 | 0.6016 | 0.5980 |
| $P<5\times10^{-5}$; $r^2<0.2$ | 219 | 142 | 0.6217 | 0.6007 | 0.5971 |
| $P<5\times10^{-6}$; - | 291 | 70 | 0.6157 | 0.5930 | 0.5946 |
| $P<5\times10^{-6}$; $r^2<0.8$ | 191 | 70 | 0.6153 | 0.5948 | 0.5946 |
| $P<5\times10^{-6}$; $r^2<0.6$ | 148 | 70 | 0.6146 | 0.5954 | 0.5948 |
| $P<5\times10^{-6}$; $r^2<0.4$ | 123 | 70 | 0.6135 | 0.5962 | 0.5939 |

| | | | | | |
|---|---|---|---|---|---|
| $P<5\times10^{-6}$; $r^2<0.2$ | 102 | 70 | 0.6138 | 0.5988 | 0.5942 |

**Table S7. The comparison of risk prediction performance for type 2 diabetes using genotype information and clinical features.** Bold numbers indicate the best performance.

| **Tuning Parameter** | **N Variants in Score** | **N Genes in Score** | **DeepPRS with clinical features AUC** | **Pruning and thresholding with clinical features AUC** | **Lasso with clinical features AUC** |
|---|---|---|---|---|---|
| $P<5\times10^{-3}$; - | 5968 | 1890 | **0.7316** | 0.7010 | 0.6953 |
| $P<5\times10^{-3}$; $r^2<0.8$ | 5069 | 1878 | 0.7306 | 0.7065 | 0.6991 |
| $P<5\times10^{-3}$; $r^2<0.6$ | 4588 | 1865 | 0.7302 | 0.7042 | 0.7018 |
| $P<5\times10^{-3}$; $r^2<0.4$ | 4203 | 1855 | 0.7291 | 0.7031 | 0.6990 |
| $P<5\times10^{-3}$; $r^2<0.2$ | 3769 | 1837 | 0.7275 | 0.6993 | 0.6944 |
| $P<5\times10^{-4}$; - | 1384 | 481 | 0.7226 | 0.7011 | **0.7053** |
| $P<5\times10^{-4}$; $r^2<0.8$ | 1059 | 478 | 0.7224 | **0.7073** | 0.7051 |
| $P<5\times10^{-4}$; $r^2<0.6$ | 919 | 477 | 0.7225 | 0.7063 | 0.7047 |
| $P<5\times10^{-4}$; $r^2<0.4$ | 816 | 475 | 0.7218 | 0.7049 | 0.7036 |
| $P<5\times10^{-4}$; $r^2<0.2$ | 700 | 472 | 0.7206 | 0.7020 | 0.7033 |
| $P<5\times10^{-5}$; - | 538 | 144 | 0.7178 | 0.7044 | 0.7033 |
| $P<5\times10^{-5}$; $r^2<0.8$ | 380 | 144 | 0.7174 | 0.7071 | 0.7045 |
| $P<5\times10^{-5}$; $r^2<0.6$ | 309 | 143 | 0.7173 | 0.7070 | 0.7038 |
| $P<5\times10^{-5}$; $r^2<0.4$ | 263 | 143 | 0.7170 | 0.7066 | 0.7041 |
| $P<5\times10^{-5}$; $r^2<0.2$ | 219 | 142 | 0.7164 | 0.7059 | 0.7031 |
| $P<5\times10^{-6}$; - | 291 | 70 | 0.7135 | 0.7036 | 0.7029 |
| $P<5\times10^{-6}$; $r^2<0.8$ | 191 | 70 | 0.7132 | 0.7043 | 0.7033 |
| $P<5\times10^{-6}$; $r^2<0.6$ | 148 | 70 | 0.7129 | 0.7044 | 0.7028 |

| | | | | | |
|---|---|---|---|---|---|
| $P<5\times10^{-6}$; $r^2<0.4$ | 123 | 70 | 0.7124 | 0.7045 | 0.7023 |
| $P<5\times10^{-6}$; $r^2<0.2$ | 102 | 70 | 0.7125 | 0.7054 | 0.7024 |

**Table S8. The comparison of risk prediction performance for breast cancer using only genotype information.** Bold numbers indicate the best performance.

| Tuning Parameter | N Variants in Score | N Genes in Score | DeepPRS AUC | Pruning and thresholding AUC | Lasso AUC |
|---|---|---|---|---|---|
| $P<5\times10^{-3}$; - | 3830 | 1553 | **0.6232** | 0.5602 | **0.6022** |
| $P<5\times10^{-3}$; $r^2<0.8$ | 3392 | 1546 | 0.6216 | 0.5551 | 0.6010 |
| $P<5\times10^{-3}$; $r^2<0.6$ | 3156 | 1544 | 0.6217 | 0.5494 | 0.6007 |
| $P<5\times10^{-3}$; $r^2<0.4$ | 2968 | 1544 | 0.6199 | 0.5437 | 0.5977 |
| $P<5\times10^{-3}$; $r^2<0.2$ | 2736 | 1537 | 0.6190 | 0.5353 | 0.5965 |
| $P<5\times10^{-4}$; - | 662 | 285 | 0.6161 | 0.5720 | 0.5980 |
| $P<5\times10^{-4}$; $r^2<0.8$ | 527 | 285 | 0.6151 | 0.5840 | 0.5982 |
| $P<5\times10^{-4}$; $r^2<0.6$ | 473 | 285 | 0.6147 | 0.5799 | 0.5976 |
| $P<5\times10^{-4}$; $r^2<0.4$ | 435 | 284 | 0.6138 | 0.5743 | 0.5952 |
| $P<5\times10^{-4}$; $r^2<0.2$ | 383 | 284 | 0.6139 | 0.5653 | 0.5944 |
| $P<5\times10^{-5}$; - | 182 | 65 | 0.6078 | **0.5920** | 0.5940 |
| $P<5\times10^{-5}$; $r^2<0.8$ | 153 | 65 | 0.6072 | 0.5903 | 0.5931 |
| $P<5\times10^{-5}$; $r^2<0.6$ | 134 | 64 | 0.6070 | 0.5882 | 0.5926 |
| $P<5\times10^{-5}$; $r^2<0.4$ | 116 | 64 | 0.6061 | 0.5856 | 0.5909 |
| $P<5\times10^{-5}$; $r^2<0.2$ | 91 | 64 | 0.6055 | 0.5820 | 0.5888 |
| $P<5\times10^{-6}$; - | 91 | 28 | 0.6004 | 0.5889 | 0.5889 |
| $P<5\times10^{-6}$; $r^2<0.8$ | 72 | 28 | 0.5997 | 0.5878 | 0.5875 |
| $P<5\times10^{-6}$; $r^2<0.6$ | 61 | 27 | 0.5996 | 0.5869 | 0.5873 |
| $P<5\times10^{-6}$; $r^2<0.4$ | 47 | 27 | 0.5988 | 0.5874 | 0.5847 |

| | | | | | |
|---|---|---|---|---|---|
| $P<5\times10^{-6}$; $r^2<0.2$ | 36 | 27 | 0.5983 | 0.5869 | 0.5839 |

**Table S9. The comparison of risk prediction performance for breast cancer using genotype information and clinical features.** Bold numbers indicate the best performance.

| Tuning Parameter | N Variants in Score | N Genes in Score | DeepPRS with clinical features AUC | Pruning and thresholding with clinical features AUC | Lasso with clinical features AUC |
|---|---|---|---|---|---|
| $P<5\times10^{-3}$; - | 3830 | 1553 | 0.6660 | 0.6286 | 0.6478 |
| $P<5\times10^{-3}$; $r^2<0.8$ | 3392 | 1546 | 0.6654 | 0.6262 | 0.6352 |
| $P<5\times10^{-3}$; $r^2<0.6$ | 3156 | 1544 | 0.6656 | 0.6238 | 0.6406 |
| $P<5\times10^{-3}$; $r^2<0.4$ | 2968 | 1544 | 0.6642 | 0.6217 | 0.6430 |
| $P<5\times10^{-3}$; $r^2<0.2$ | 2736 | 1537 | 0.6636 | 0.6189 | 0.6444 |
| $P<5\times10^{-4}$; - | 662 | 285 | 0.6622 | 0.6350 | 0.6485 |
| $P<5\times10^{-4}$; $r^2<0.8$ | 527 | 285 | 0.6612 | 0.6422 | 0.6495 |
| $P<5\times10^{-4}$; $r^2<0.6$ | 473 | 285 | 0.6609 | 0.6398 | 0.6482 |
| $P<5\times10^{-4}$; $r^2<0.4$ | 435 | 284 | 0.6604 | 0.6366 | 0.6472 |
| $P<5\times10^{-4}$; $r^2<0.2$ | 383 | 284 | 0.6604 | 0.6318 | 0.6475 |
| $P<5\times10^{-5}$; - | 182 | 65 | 0.6566 | 0.6467 | 0.6480 |
| $P<5\times10^{-5}$; $r^2<0.8$ | 153 | 65 | 0.6558 | 0.6454 | 0.6466 |
| $P<5\times10^{-5}$; $r^2<0.6$ | 134 | 64 | 0.6561 | 0.6442 | 0.6467 |
| $P<5\times10^{-5}$; $r^2<0.4$ | 116 | 64 | 0.6554 | 0.6425 | 0.6455 |
| $P<5\times10^{-5}$; $r^2<0.2$ | 91 | 64 | 0.6551 | 0.6401 | 0.6439 |
| $P<5\times10^{-6}$; - | 91 | 28 | 0.6517 | 0.6452 | 0.6448 |
| $P<5\times10^{-6}$; $r^2<0.8$ | 72 | 28 | 0.6513 | 0.6445 | 0.6435 |
| $P<5\times10^{-6}$; $r^2<0.6$ | 61 | 27 | 0.6515 | 0.6441 | 0.6433 |

| | | | | | |
|---|---|---|---|---|---|
| $P<5\times10^{-6}$; $r^2<0.4$ | 47 | 27 | 0.6507 | 0.6443 | 0.6419 |
| $P<5\times10^{-6}$; $r^2<0.2$ | 36 | 27 | 0.6505 | 0.6440 | 0.6414 |

**Table S10. Cases with few known risk SNPs but high deep polygenic risk scores.** We define the SNPs with *P* value less than 5e-8 and the effect estimate (*β*) greater than 0 in GWAS as significant risk SNPs.

| **Sample ID** | **Risk SNPs included in GWAS (n)** | **Mutations in Risk SNPs (n)** | **Pruning and thresholding method percentile score** | **DeepPRS percentile score** | **Odds ratio based on DeepPRS** |
|---|---|---|---|---|---|
| **Alzheimer's disease** | | | | | |
| Individual 1 | 36 | 5 | 1 | 99 | 9.66 |
| Individual 2 | 36 | 5 | 1 | 99 | 9.53 |
| Individual 3 | 36 | 6 | 1 | 99 | 10.59 |
| **Inflammatory bowel disease** | | | | | |
| Individual 4 | 236 | 36 | 5 | 99 | 4.23 |
| Individual 5 | 236 | 42 | 5 | 99 | 4.64 |
| Individual 6 | 236 | 46 | 6 | 99 | 5.21 |
| **Type 2 diabetes** | | | | | |
| Individual 7 | 56 | 8 | 1 | 75 | 2.34 |
| Individual 8 | 56 | 7 | 2 | 98 | 3.27 |
| Individual 9 | 56 | 8 | 9 | 86 | 2.50 |
| **Breast cancer** | | | | | |
| Individual 10 | 23 | 2 | 5 | 89 | 2.23 |
| Individual 11 | 23 | 3 | 19 | 91 | 2.29 |
| Individual 12 | 23 | 3 | 19 | 97 | 2.67 |

**Table S11. An illustration example of calculating odds ratio.** Odds ratios are calculated by comparing those with high deep polygenic risk score to the remainder of the population. OR= $(\mathbf{T}_{\boldsymbol{D}}\mathbf{R}_{\boldsymbol{E}})/(\mathbf{T}_{\boldsymbol{E}}\mathbf{R}_{\boldsymbol{D}})$.

| Population/Phenotype | Diseased | Healthy |
|---|---|---|
| Top 20% of distribution | $T_D$ | $T_E$ |
| Remaining 80% | $R_D$ | $R_E$ |
